# Feasibility and Measurement Properties of a Large Language Model–Based Virtual Patient Laboratory for Clinical Reasoning Training in Physical Therapy Students: Single-Arm Pilot Study

**DOI:** 10.64898/2026.10.13.26364741

**Authors:** Alfredo Lerín-Calvo, Alberto Pérez-González, Asier Sagarribay Villano, Silvia Di-Bonaventura, Raúl Ferrer-Peña

## Abstract

**Background:** Large language models (LLMs) configured as virtual patients may expand structured practice of clinical reasoning in health professions education. Before testing effectiveness, it must be established whether students engage with them and whether the resulting scores are complete, reliable, and related to performance with real patients. Our earlier pilot randomized trial found very low engagement with a less structured LLM-based approach.

**Objective:** This study aimed to assess the feasibility (adherence, acceptability, and data completeness) of a revised, structured LLM-based clinical reasoning laboratory (Laboratorio de Inteligencia Artificial [LIA]) in undergraduate physical therapy education and to explore its measurement properties, including its association with clinical judgment with a real patient assessed with the Lasater Clinical Judgment Rubric (LCJR).

**Methods:** In this single-arm pilot feasibility study, 27 third-year physical therapy students at a Spanish university completed, in 2024-2025, 3 virtual patient cases (lumbar, cervical, and thoracic spine pain) simulated by ChatGPT (GPT-4) over 4 weeks, each twice. ChatGPT scored each attempt from 0 to 4 on the same 10 dimensions of the Lasater Clinical Judgment Rubric (LCJR; 0-40 points) that human assessors applied (1-4 per dimension; 10-40 points) during a real patient encounter. Feasibility outcomes were case completion, data completeness, and satisfaction (0-10). Measurement outcomes were internal consistency and convergent validity (Spearman correlations between model-generated and human LCJR scores). Variability estimates informed the sample size of a future trial.

**Results:** All 27 students were assessed with the LCJR; 22 (81%) completed at least 1 LIA case. Both attempts of the lumbar, cervical, and thoracic cases were completed by 78%, 63%, and 74% of students, respectively. Satisfaction data were available for 15 students (56%; mean 6.0, SD 1.3), and missing secondary data clustered in the third rotation. Internal consistency was excellent for the LCJR (α=.96) and ranged from .43 to .90 for model-generated ratings. Model-generated LIA scores clustered near the maximum (mean 36.0, SD 1.4). Among completers, LIA scores correlated weakly with the LCJR (ρ=0.25, 95% CI –0.20 to 0.61; ρ=0.44, 95% CI 0.05-0.71, when noncompleted attempts were scored as 0). Grade point average correlated almost perfectly with the LCJR (ρ=0.96; n=22); after adjustment for it, the LIA-LCJR association disappeared (partial ρ=–0.10). A future trial would require approximately 39 students per group (80% power, conservative variance).

**Conclusions:** Delivering the LIA within the curriculum was feasible, but clinical judgment ratings generated by ChatGPT did not reflect those of human assessors with a real patient: they clustered near the maximum and showed no convergent validity once noncompletion was separated from performance. LLM-generated ratings should not be used as outcome or assessment measures without validation against human raters. Case calibration, completion tracking, and clarification of the GPA-LCJR relationship are also needed before a definitive trial.

**Trial Registration:** ClinicalTrials.gov NCT06809634; https://clinicaltrials.gov/study/NCT06809634

## Introduction

Clinical reasoning is a core competency in physical therapy, but providing students with repeated, structured, and feedback-rich opportunities to practice it remains challenging. Conversational artificial intelligence is increasingly being considered as a way to expand opportunities for clinical reasoning practice in health professions education [1]. In physical therapy, this interest is particularly relevant because students often need repeated exposure to clinical scenarios before they can confidently manage the uncertainty and interactional demands of real patient encounters [2].

Current approaches to clinical reasoning training have practical limitations. Clinical placements provide authentic experience, but the cases, feedback, and learning opportunities available to each student vary substantially across settings [3]. Simulation with standardized patients offers a more controlled alternative, yet it requires trained actors or facilitators, physical space, coordination, and faculty time [4]. These constraints limit the number of scenarios students can complete and the amount of individualized feedback they receive.

Large language model (LLM)–based virtual patients may help address some of these constraints by providing structured, repeatable, and adaptable clinical encounters at low marginal cost [5,6]. When designed around fixed cases and predefined scoring criteria, these tools may allow students to practice history taking, hypothesis generation, clinical interpretation, and treatment planning in a controlled digital environment. However, their feasibility cannot be assumed. LLM outputs vary with prompting, may be inconsistent across interactions, and may occasionally be inaccurate or implausible [7,8]. Engagement is a further concern: digital educational tools tend to be underused when they are not clearly integrated into coursework, perceived as useful, or supported by appropriate guidance [9,10]. Partial completion and missing data compromise both educational value and research interpretation, so feasibility studies should examine not only satisfaction but also whether students complete the activities, whether data can be collected consistently, and whether outcome measures are suitable for a future trial [11].

In a previous pilot randomized trial, we allocated third-year physical therapy students to a 4-week LLM-based virtual patient program or to usual placement-based training [12]. Recruitment was complete, but only 4 of the 23 students allocated to the LLM arm engaged actively with the model, and we concluded that preparatory training and closer curricular integration were needed before a definitive trial. That study did not examine the measurement properties of the LLM-based performance scores, their relationship with an independent real-patient assessment of clinical judgment, or the pattern of missing data across a full cohort. Evidence on these questions in physical therapy remains scarce [10], and they must be answered before a comparative study can attribute educational effects to the intervention.

This study therefore examined the feasibility of a revised, structured LLM-based clinical reasoning laboratory for undergraduate physical therapy students. The objectives were to assess (1) adherence and case completion, (2) acceptability, (3) completeness and pattern of missing data, and (4) the suitability of the outcome measures, including internal consistency and preliminary convergent validity against clinical judgment assessed with the Lasater Clinical Judgment Rubric (LCJR) during a real patient encounter [13,14], and (5) to obtain variability estimates to inform the sample size of a future randomized trial.

## Methods

### Study Design

This was a single-arm pilot feasibility study guided by the framework of Bowen et al [15], which distinguishes acceptability, demand, implementation, practicality, and related domains as the core questions that determine whether an intervention is ready for a definitive trial. The design is consistent with previous feasibility studies in physical therapy [16,17]. The students included here constituted the intervention arm of a larger quasi-experimental study conducted in the same academic year; the comparison between arms is reported in a separate manuscript (Lerín-Calvo A, et al, unpublished data) and is outside the scope of this report. The present analysis addresses questions that do not require a comparison group: whether the intervention can be delivered and measured adequately.

The study is reported following the CONSORT extension for pilot and feasibility trials [18], adapted for a single-arm design, and the CONSORT-EHEALTH checklist [19] (Multimedia Appendix 1), and it follows consolidated guidance for pilot and feasibility studies [11]. No quantitative progression criteria were specified a priori; feasibility outcomes are therefore interpreted descriptively, and explicit criteria are proposed for the next stage (see Discussion).

### Ethical Considerations

The study was approved by the ethics committee of the La Salle University Center for Advanced Studies, Madrid, Spain (CSEULS-PI-002/2025), and the study was registered previously in ClinicalTrials.gov (NCT06809634). All information and consent materials were provided to participants in an easy-read format, and written informed consent was obtained from every participant before enrollment. Participation was voluntary, and the decision to participate or to withdraw had no effect on course grades. Students were instructed not to enter personal data into the LLM, and virtual patient cases were fictitious; study data were pseudonymized before analysis and stored on institutional servers with access restricted to the research team. Participants received no financial compensation.

### Participants and Setting

Participants were undergraduate physical therapy students at Centro Superior de Estudios Universitarios La Salle, Universidad Autónoma de Madrid (Spain), who received the LLM-based clinical reasoning intervention during the 2024-2025 academic year. Students were eligible if they were enrolled in the third year of the physical therapy degree, were taking Practicum I, a clinical placement course, during the 2024-2025 academic year, and provided written informed consent. No exclusion criteria were applied other than withdrawal of consent.

The course is organized in 3 sequential clinical rotations; students completed the LIA during the rotation to which they were assigned (first rotation, n=8; second, n=8; third, n=11). As is customary in feasibility research, no a priori sample size calculation was performed [20]; the sample comprised all eligible students in the intervention arm.

### Intervention: LLM-Based Clinical Reasoning Laboratory

The LIA consisted of structured interactions with ChatGPT (OpenAI; GPT-4), used between September 2024 and January 2025 and configured through predefined prompts to act as a standardized virtual patient. The prompt structure was adapted from our previous pilot trial [12]; the configuration prompt and an example interaction for the lumbar case are provided in Multimedia Appendix 2. Three fictitious cases were developed by one of the authors (RFP), a physical therapist with 15 years of clinical experience: (1) lumbar pain of facet joint origin, (2) cervical pain, and (3) thoracic spine pain. Each student completed each case twice, yielding up to 6 case attempts per student. As in the earlier pilot trial [12], the program lasted 4 weeks. As in that trial, the predefined prompts were provided in a PDF file, each student used a personal ChatGPT account, the cases were completed autonomously without faculty supervision of the interactions, and no formal training in LLM use was provided beforehand.

Compared with the previous pilot trial [12], in which students completed 4 clinically heterogeneous cases, 1 per week, and engagement was very low, the program was restructured into 3 spine cases, each completed twice.

At the end of each attempt, students asked ChatGPT, through predefined prompts, to evaluate their reasoning in the conversation by applying the LCJR and to score each dimension (Multimedia Appendix 2). The model applied the same 10-dimension version of the LCJR used for the real patient assessment (Multimedia Appendix 3), scoring each dimension from 0 to 4 (4 = exemplary), for a total of 0 to 40 points per attempt. This 5-point scale did not match the 4 performance levels used by human assessors (1-4). Observed dimension scores ranged from 2 to 4. The model-generated dimension scores were recorded in the study database; no human rater scored the attempts.

### Outcome Measures

#### Feasibility Outcomes

Following Bowen et al [15] and previous physical therapy feasibility studies [16,17], 4 feasibility domains were assessed. Adherence was defined as the proportion of students who completed each case attempt; an attempt was considered completed when a rubric score generated by the model was recorded for it; attempts without a recorded score, or recorded as 0, were considered not completed. Data completeness was the proportion of students with available data for each measure, examined overall and by clinical rotation. Acceptability was assessed with satisfaction with the educational program, rated on a 0 to 10 visual analog scale at the end of the intervention. Suitability of the outcome measures was assessed through their score distributions (floor and ceiling effects), internal consistency, and, for the LIA, convergent validity.

#### LIA Performance

For each student, the LIA summary score was the mean total score across all completed case attempts (possible range 0-40). The best-attempt score for each case and the change between the first and second attempts within each case were also derived.

#### Clinical Judgment With a Real Patient (Convergent Validity Criterion)

Clinical judgment was assessed with the LCJR [13], which comprises 11 behavioral dimensions grouped into 4 phases (noticing, interpreting, responding, and reflecting) scored on 4 levels. In this study, a 10-dimension version was used (total score 10-40), with 1 dimension in the reflecting phase (commitment to improvement); the evaluation/self-analysis dimension of the original rubric was omitted (Multimedia Appendix 3) because it requires a structured debriefing after the encounter, which was not part of the real patient assessment. We used the Spanish version culturally adapted and validated in nursing students [21], replacing references to nursing with physical therapy. The LCJR was scored by the instructor of the Practicum I course during a real patient encounter in the clinical placement at the end of the 4-week program; the instructor was not aware of students’ LIA scores. The LCJR was developed and validated mainly in nursing education [13,22]. Because the same rubric was applied by ChatGPT in the virtual cases and by human assessors with a real patient, LIA scores were expected to correlate positively with LCJR scores if model-generated ratings captured students’ clinical judgment.

#### Secondary Measures

Digital competencies were assessed with a 26-item questionnaire covering 6 domains of digital competence for health care professionals [23], the instrument used in our previous pilot [12]; the total score was the sum of all items. Self-evaluation of clinical competence was rated on a single item (1-4 scale). Academic grade point average (GPA; 0-10) and age were obtained from academic records; GPA did not include the clinical placement grade.

### Statistical Analysis

Missing data were quantified by variable and by clinical rotation and inspected graphically. Continuous variables were summarized as mean (SD), median, and range, and proportions with Wilson 95% CIs. Internal consistency was estimated with Cronbach α for the LCJR, the digital competencies questionnaire, and the model-generated LCJR ratings (LIA scores), the latter separately for each case attempt among students who completed it. Agreement between first and second attempts within each case was estimated with the intraclass correlation coefficient (ICC; 2-way mixed, consistency, single measures).

Convergent validity was estimated with Spearman correlations between the LIA summary score and the LCJR total and phase scores, with 95% CIs obtained with the Bonett-Wright method [24]. The primary analysis included students with at least 1 completed case attempt, with noncompleted attempts treated as missing. In a sensitivity analysis, noncompleted attempts were scored as 0, as in the original data coding. Because academic performance may influence both measures, the LIA-LCJR association was additionally estimated as a partial Spearman correlation adjusted for GPA among students with available GPA. LCJR scores of students who did and did not complete the LIA were compared with the Mann-Whitney U test.

Changes between attempts were examined with Wilcoxon signed rank tests among students who completed both attempts of each case. Spearman correlations were also computed among all instruments (Multimedia Appendix 4). To inform a future 2-arm randomized trial, SDs with 95% CIs were estimated for candidate primary outcomes, and sample sizes were calculated for a 2-tailed independent-samples t test (α=.05; power 80% and 90%) using both the observed SD and the upper limit of its one-sided 80% CI, as recommended for pilot-derived estimates [25]. Target differences corresponded to approximately 0.7 SD of the observed distributions (eg, 5 points for the LCJR), because no minimal important difference has been established for these measures in physical therapy students.

Given the feasibility purpose and small sample, analyses were interpreted descriptively, with emphasis on estimates and their precision rather than hypothesis testing; no correction for multiple comparisons was applied, and P values are reported for completeness only. Analyses were conducted in R (version 4.3.3; R Foundation for Statistical Computing) with the dplyr, tidyr, and ggplot2 packages.

## Results

### Participants and Data Completeness

The 27 students (10 of the 22 with available data were women; mean age 22.5 years, SD 3.2; n=22 with available age) were distributed across the first (n=8), second (n=8), and third (n=11) rotations (Table 1). LCJR scores were available for all 27 students. Data were available for self-evaluation in 24 students (89%), GPA in 22 (81%), digital competencies in 21 (78%), and satisfaction in 15 (56%) (Table 2, Figure 1). Missing data were concentrated in the third rotation, in which all 11 students lacked at least 1 secondary measure, compared with 2 of 16 in the first 2 rotations, which indicates a problem in the data collection procedure during that period rather than random individual nonresponse.

**Figure 1.**
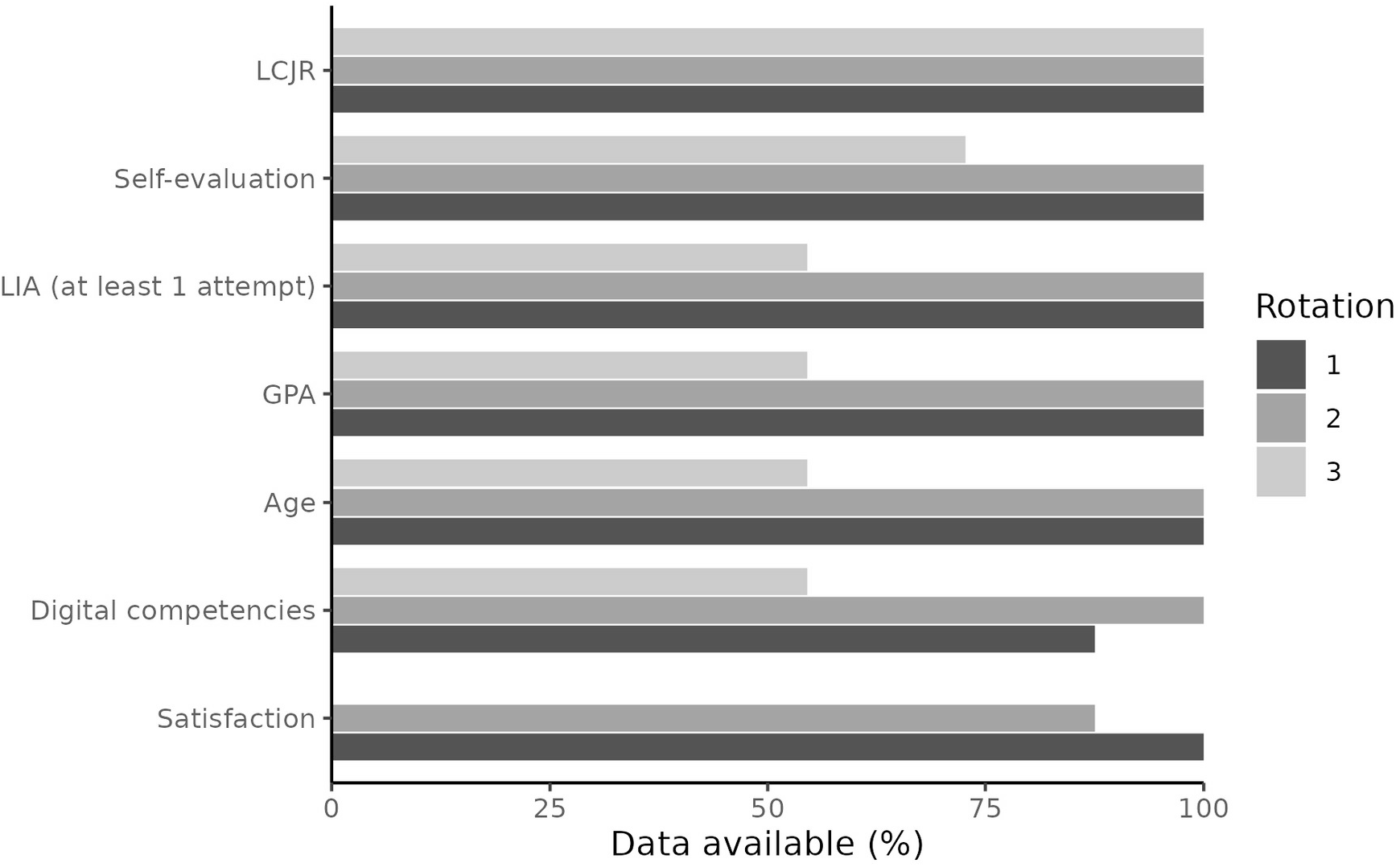
Availability of data by variable and clinical rotation (n=27).

**Table 1.** Participant characteristics and data availability by clinical rotation.

| Characteristic | Overall (n=27) | Rotation 1 (n=8) | Rotation 2 (n=8) | Rotation 3 (n=11) |
| --- | --- | --- | --- | --- |
| Women, n (%) | 10/22 (45) | 5/8 (63) | 3/8 (38) | 2/6 (33) |
| Age (years), mean (SD)<sup>a</sup> | 22.5 (3.2) | 21.4 (1.2) | 22.1 (2.3) | 24.5 (5.1) |
| GPA (0-10), mean (SD)<sup>b</sup> | 8.45 (1.32) | 8.12 (1.60) | 9.06 (0.44) | 8.06 (1.59) |
| LCJR available, n (%) | 27 (100) | 8 (100) | 8 (100) | 11 (100) |
| ≥1 LIA attempt completed, n (%) | 22 (81) | 8 (100) | 8 (100) | 6 (55) |
| Digital competencies available, n (%) | 21 (78) | 7 (88) | 8 (100) | 6 (55) |
| Self-evaluation available, n (%) | 24 (89) | 8 (100) | 8 (100) | 8 (73) |
| Satisfaction available, n (%) | 15 (56) | 8 (100) | 7 (88) | 0 (0) |
| GPA available, n (%) | 22 (81) | 8 (100) | 8 (100) | 6 (55) |

<sup>a</sup>Available for 22 students (6 in rotation 3). <sup>b</sup>Available for 22 students (6 in rotation 3). Sex was available for 22 students (6 in rotation 3). GPA: grade point average; LCJR: Lasater Clinical Judgment Rubric; LIA: Laboratorio de Inteligencia Artificial (artificial intelligence–based clinical reasoning laboratory).

**Table 2.** Feasibility outcomes: adherence and data completeness (N=27).

| Indicator | n/N | % (95% CI)<sup>a</sup> |
| --- | --- | --- |
| Assessed with the LCJR | 27/27 | 100 (88-100) |
| Completed ≥1 LIA case attempt | 22/27 | 81 (63-92) |
| Completed all 6 LIA case attempts | 14/27 | 52 (34-69) |
| Lumbar case, both attempts completed<sup>b</sup> | 21/27 | 78 (59-89) |
| Cervical case, both attempts completed<sup>b</sup> | 17/27 | 63 (44-78) |
| Thoracic case, both attempts completed<sup>b</sup> | 20/27 | 74 (55-87) |
| Self-evaluation available | 24/27 | 89 (72-96) |
| GPA available | 22/27 | 81 (63-92) |
| Digital competencies available | 21/27 | 78 (59-89) |
| Satisfaction available | 15/27 | 56 (37-72) |

<sup>a</sup>Wilson score interval. <sup>b</sup>Attempts without a recorded score, or recorded as 0, were considered not completed. GPA: grade point average; LCJR: Lasater Clinical Judgment Rubric; LIA: Laboratorio de Inteligencia Artificial (artificial intelligence-based clinical reasoning laboratory).

### Feasibility Outcomes

#### Adherence

Overall, 22 of 27 students (81%) completed at least 1 LIA case attempt and 14 (52%) completed all 6 attempts; 5 students, all in the third rotation, completed none. Both attempts were completed by 21 of 27 students (78%, 95% CI 59-89) for the lumbar case, 17 of 27 (63%, 95% CI 44-78) for the cervical case, and 20 of 27 (74%, 95% CI 55-87) for the thoracic case (Table 2).

#### Acceptability

Satisfaction data were available for 15 of 27 students (56%), none of them in the third rotation. Mean satisfaction was 6.0 (SD 1.3; median 6.0; range 4-8) on the 0 to 10 scale. No adverse events or unintended effects were identified.

### Score Distributions and Internal Consistency

Descriptive statistics and internal consistency are shown in Table 3. LCJR scores were negatively skewed (median 34.0; range 16-40), with a subgroup of 3 students scoring below 20. Internal consistency was excellent for the LCJR (α=.96) and good for the digital competencies questionnaire (α=.90). For the model-generated ratings, α ranged from .43 to .90 across case attempts, with the lowest values for the cervical case. LIA scores were concentrated near the top of the scale: the LIA summary score ranged from 32.7 to 37.8 (mean 36.0, SD 1.4), and the cervical case showed the most marked ceiling effect (first attempt mean 37.6 of 40, SD 1.7), which restricts variance and lowers any reliability or validity estimate. Agreement between first and second attempts was low: ICC 0.35 (lumbar), 0.35 (cervical), and –0.12 (thoracic) (Table 5).

**Table 3.** Descriptive statistics and internal consistency of outcome measures.

| Instrument (possible range) | n | Mean (SD) | Median | Range | Cronbach $\alpha$ |
| --- | --- | --- | --- | --- | --- |
| LCJR total (10-40) | 27 | 31.44 (6.76) | 34.0 | 16-40 | .96 |
| Noticing (3-12) | 27 | 9.59 (1.78) | 10.0 | 6-12 | — |
| Interpreting (2-8) | 27 | 5.93 (1.33) | 6.0 | 3-8 | — |
| Responding (4-16) | 27 | 12.52 (3.26) | 14.0 | 4-16 | — |
| Reflecting (1-4) | 27 | 3.41 (0.84) | 4.0 | 1-4 | — |
| LIA summary score (0-40)<sup>a</sup> | 22 | 35.98 (1.38) | 36.5 | 32.7-37.8 | .43 to .90 (by case attempt) |
| Digital competencies total (26 items) | 21 | 159.95 (22.37) | 157.0 | 126-213 | .90 |
| Self-evaluation (1-4) | 24 | 3.12 (0.54) | 3.0 | 2-4 | — |
| Satisfaction (0-10) | 15 | 6.00 (1.30) | 6.0 | 4-8 | — |
| GPA (0-10) | 22 | 8.45 (1.32) | 9.0 | 5-10 | — |

<sup>a</sup>Mean of completed case attempts. With noncompleted attempts coded as 0 (original coding), n=27, mean 26.43 (SD 13.68), median 32.7, range 0-37.8. GPA: grade point average; LCJR: Lasater Clinical Judgment Rubric; LIA: Laboratorio de Inteligencia Artificial (artificial intelligence–based clinical reasoning laboratory).

### Convergent Validity

Among the 22 students who completed at least 1 LIA case attempt, the LIA summary score correlated weakly and imprecisely with the LCJR total (ρ=0.25, 95% CI –0.20 to 0.61) and with the responding phase (ρ=0.14, 95% CI –0.30 to 0.53) (Table 4, Figure 2). When noncompleted attempts were scored as 0, the corresponding correlations were ρ=0.44 (95% CI 0.05-0.71) and ρ=0.32 (95% CI –0.08 to 0.63). Students who completed no LIA attempt had lower LCJR scores than those who did (median 30.0 vs 34.0; P=.07), so the larger association in the sensitivity analysis largely reflects engagement rather than performance. After adjustment for GPA, the partial correlation between LIA and LCJR was ρ=–0.10 (95% CI –0.51 to 0.35; n=22).

**Figure 2.**
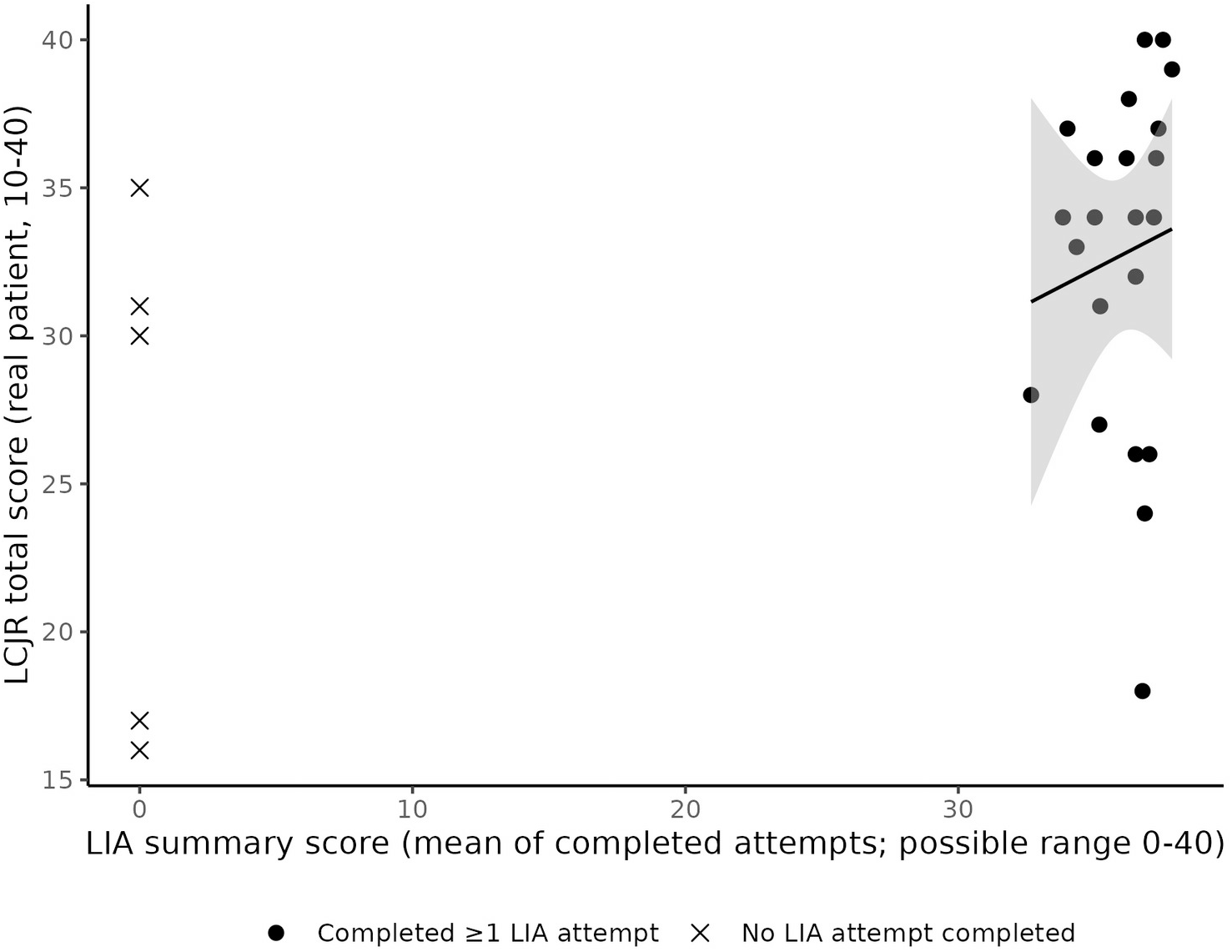
Relationship between the LIA summary score and the LCJR total score. Students who completed no LIA attempt are shown separately and are excluded from the primary analysis.

**Table 4.**
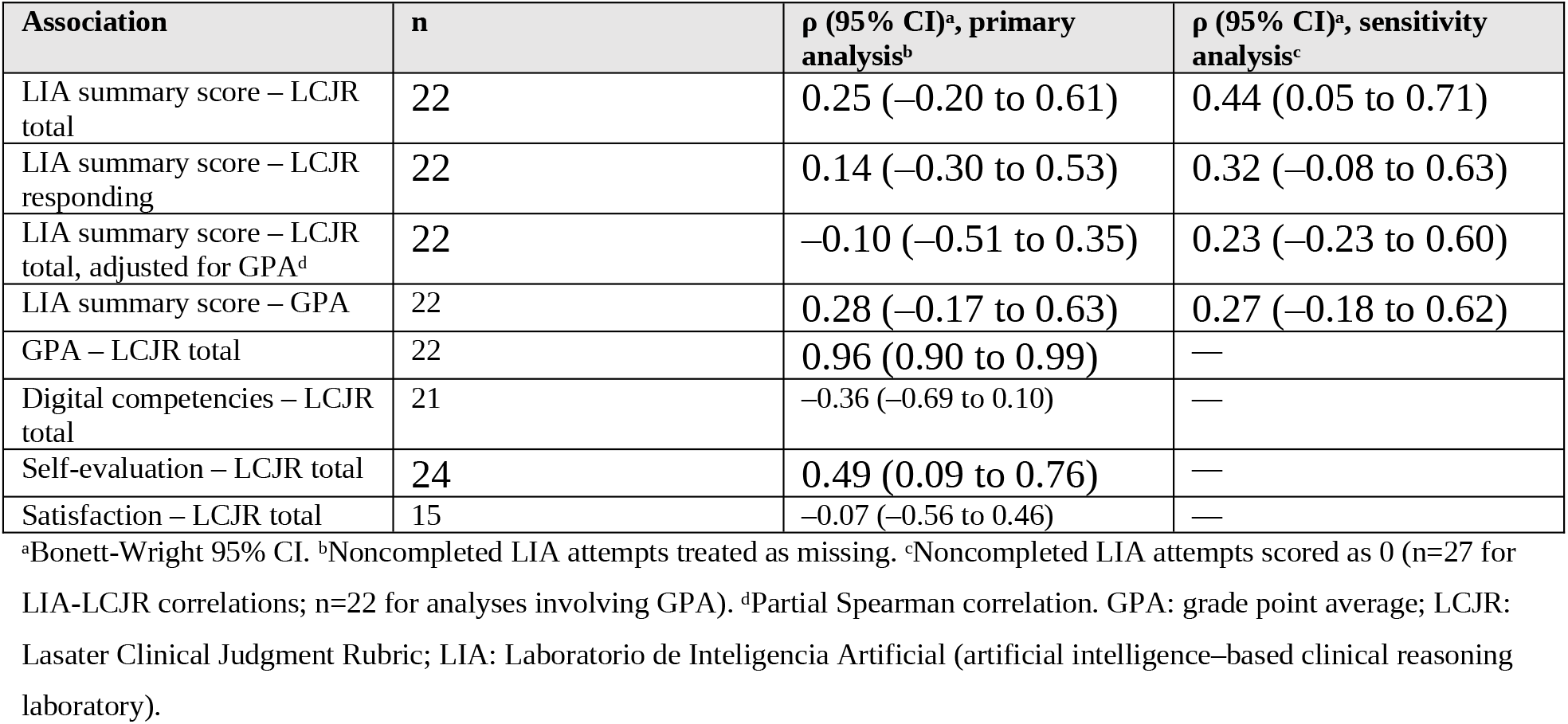
Spearman correlations with clinical judgment (LCJR).

**Table 5.** LIA scores on the first and second attempts among students who completed both attempts of each case.

| Case | n | Attempt 1, mean (SD) | Attempt 2, mean (SD) | Change, mean (SD) | ICC (95% CI) | P value<sup>a</sup> |
| --- | --- | --- | --- | --- | --- | --- |
| Lumbar | 21 | 35.0 (3.0) | 35.8 (3.3) | 0.8 (3.6) | 0.35 (−0.09 to 0.67) | .29 |
| Cervical | 17 | 37.4 (1.7) | 37.8 (2.1) | 0.4 (2.2) | 0.35 (−0.15 to 0.70) | .38 |
| Thoracic | 20 | 34.4 (4.3) | 36.0 (4.1) | 1.6 (6.3) | −0.12 (−0.53 to 0.33) | .25 |

<sup>a</sup>Wilcoxon signed rank test. ICC: intraclass correlation coefficient (2-way mixed, consistency, single measures); LIA: Laboratorio de Inteligencia Artificial (artificial intelligence–based clinical reasoning laboratory).

GPA showed an almost perfect correlation with the LCJR (ρ=0.96, 95% CI 0.90-0.99; n=22) and a weak correlation with the LIA summary score (ρ=0.28, 95% CI –0.17 to 0.63). Self-evaluation showed a moderate positive correlation with the LCJR (ρ=0.49, 95% CI 0.09-0.76; n=24), whereas digital competencies (ρ=–0.36, 95% CI –0.69 to 0.10; n=21) and satisfaction (ρ=–0.07, 95% CI –0.56 to 0.46; n=15) were not clearly associated with it. The full correlation matrix is provided in Multimedia Appendix 4.

### Change Between Attempts

No case showed a meaningful change between the first and second attempts among students who completed both (Table 5, Figure 3). Mean changes were 0.8 points (SD 3.6) for the lumbar case, 0.4 points (SD 2.2) for the cervical case, whose first-attempt scores were already close to the maximum, and 1.6 points (SD 6.3) for the thoracic case.

**Figure 3.**
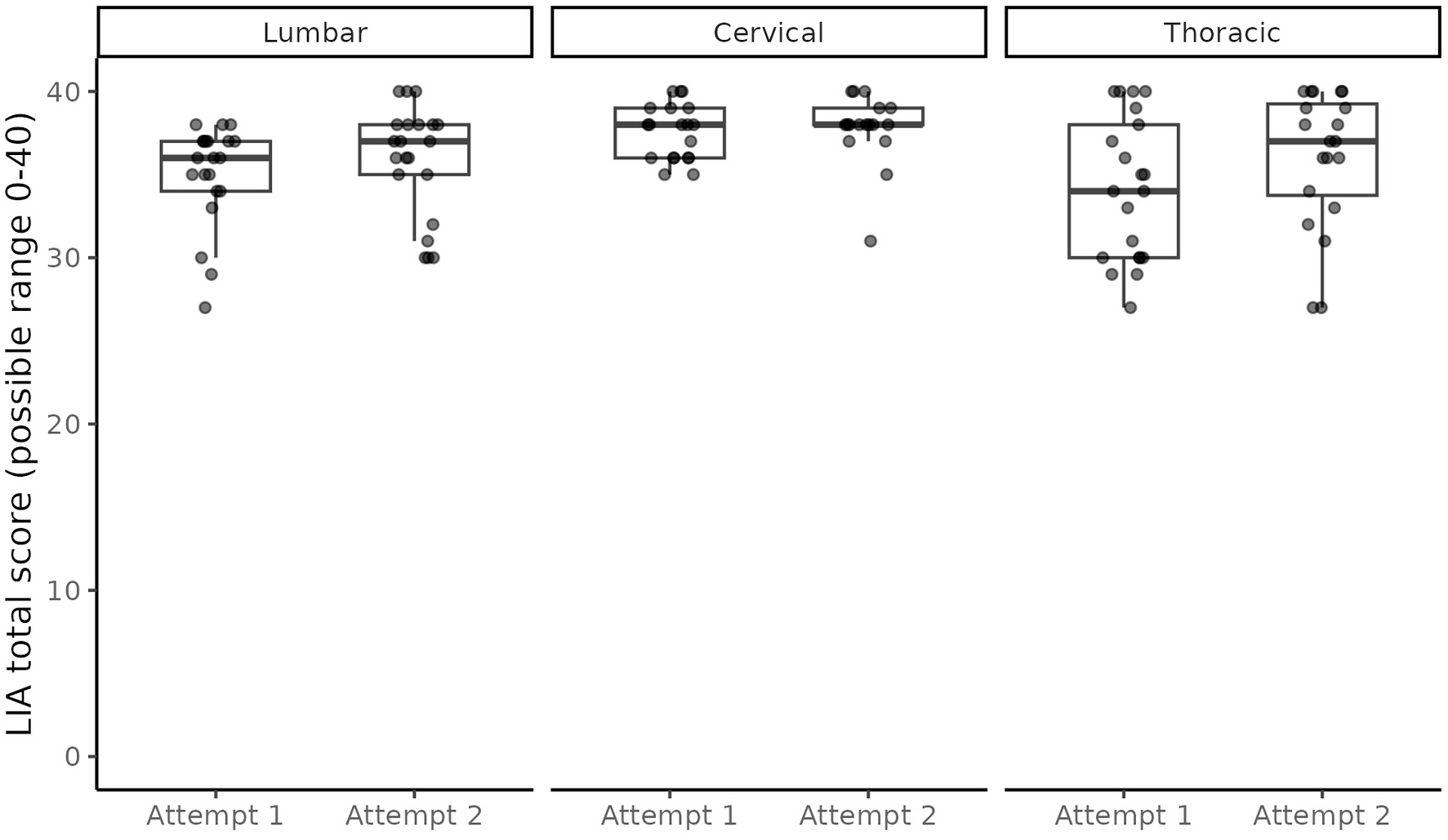
LIA total scores by case and attempt among students who completed each attempt.

### Variability Estimates for a Future Randomized Trial

Variability estimates and projected sample sizes are shown in Table 6. For the LCJR total, the responding phase, and digital competencies, the observed SDs yielded 28 to 36 students per group for 80% power to detect differences of about 0.7 SD; using the upper limit of the one-sided 80% CI of the SD, 37 to 49 students per group would be required (48 to 66 for 90% power). The SD of the LIA summary score was very small (1.4) because of the ceiling effect, so sample size estimates based on it are not informative until the scoring procedure is revised. For the LCJR, a difference of 5 points would require 39 students per group under the conservative estimate.

**Table 6.** Variability estimates and projected sample size per group for a future 2-arm randomized trial (2-tailed α=.05).

| Outcome | n | Mean | SD (95% CI) | Target difference (SD units) | n/group, 80% power<sup>a</sup> | n/group, 90% power<sup>a</sup> | n/group, 80% power<sup>b</sup> | n/group, 90% power<sup>b</sup> |
| --- | --- | --- | --- | --- | --- | --- | --- | --- |
| LCJR total | 27 | 31.44 | 6.76 (5.32-9.27) | 5 (0.74) | 30 | 40 | 39 | 52 |
| LCJR responding | 27 | 12.52 | 3.26 (2.57-4.47) | 2.5 (0.77) | 28 | 37 | 37 | 48 |
| Digital competencies | 21 | 159.95 | 22.37 (17.11-32.30) | 15 (0.67) | 36 | 48 | 49 | 66 |
| LIA summary score | 22 | 35.98 | 1.38 (1.06-1.97) | 1.0 (0.72) | 31 | 42 | 42 | 56 |

<sup>a</sup>Based on the observed SD. <sup>b</sup>Based on the upper limit of the one-sided 80% CI of the SD [25]. Sample sizes do not include allowance for attrition. LCJR: Lasater Clinical Judgment Rubric; LIA: Laboratorio de Inteligencia Artificial (artificial intelligence-based clinical reasoning laboratory).

## Discussion

### Principal Findings

This pilot study assessed the feasibility of integrating a structured LLM-based clinical reasoning laboratory into undergraduate physical therapy education and the validity of the scores generated within it. The main finding concerns validity: when ChatGPT applied the same clinical judgment rubric that human assessors used with a real patient, its ratings clustered near the maximum, correlated only weakly with the human ratings among students who completed the cases, and were unrelated to them after adjustment for GPA. The apparent association observed when noncompleted attempts were scored as 0 reflected engagement rather than performance. In contrast, the intervention could be delivered within the curriculum with higher engagement than in our previous pilot trial, although completion varied across cases, missing secondary data clustered in 1 rotation, and GPA correlated almost perfectly with the LCJR.

### Lack of Convergent Validity of Model-Generated Ratings

Because the same 10-dimension LCJR was applied by ChatGPT in the virtual cases and by human assessors with a real patient, the 2 sets of scores differed mainly in the rater and in the context of the encounter, which makes their lack of agreement informative. Several mechanisms may explain it. First, the model-generated ratings were lenient and compressed: the LIA summary score ranged from 32.7 to 37.8 of 40 (SD 1.4), whereas human LCJR scores ranged from 16 to 40, so the model barely discriminated between students. In addition, the model was asked to score each dimension from 0 to 4, a 5-point scale that did not match the 4 performance levels of the rubric applied by human assessors, so the 2 sets of scores were not on exactly the same metric; nevertheless, the model never assigned a score below 2 to any dimension, and the lower part of its scale was never used. Leniency is plausible because the rating was requested by the student at the end of a cooperative conversation, a setting in which general-purpose LLMs tend to produce agreeable, positive feedback, and because the model rated only what the student typed, without access to examination skills, nonverbal communication, or the patient’s response, which are central to the noticing and responding phases of the LCJR. Second, the ratings were unstable: agreement between the 2 attempts of the same case was poor (ICC –0.12 to 0.35) even though mean scores barely changed, consistent with evidence that ChatGPT responses vary across repeated queries [7,8]. Third, internal consistency of the model-generated ratings was low for the cervical case (α=.43-.67), whose scores were closest to the ceiling. Fourth, the association observed when noncompleted attempts were scored as 0 (ρ=0.44) was largely an artifact of that coding: students who did not engage with the LIA tended to have lower clinical judgment scores (median 30.0 vs 34.0), so the correlation captured engagement rather than performance.

These findings have practical implications. Emerging evidence suggests that LLM-based simulations can capture meaningful aspects of clinical performance in other health professions [1,26], and LLM-generated feedback is increasingly proposed as a scalable form of assessment. Our data indicate that, at least when the model rates a student-requested evaluation through a general-purpose prompt, LLM-generated scores should not be used as outcome measures, for grading, or as evidence of learning without prior validation against trained human raters. Such validation should examine agreement with human ratings of the same transcripts, use explicit behavioral anchors for each rubric level, and keep both the model and human raters unaware of other information about the student. The human LCJR also requires scrutiny: it showed excellent internal consistency, in line with previous reports in nursing [22], although α values above .95 may indicate item redundancy or a global rating tendency, and it correlated almost perfectly with GPA (ρ=0.96). Academic performance is a recognized predictor of clinical performance [27], but a correlation this close to unity between a grade average and a rubric-based observation of a single encounter is unusual. Because GPA excluded the placement grade, it cannot be explained by the LCJR contributing to GPA; it may instead reflect the rater’s knowledge of students’ academic performance, as the LCJR was scored by the course instructor, or chance in a subsample of 22 students. Until this is clarified, GPA should be treated as a covariate and the LCJR should be scored by raters independent of course grading. Finally, the absence of improvement between attempts should not be interpreted as evidence against learning, because ceiling-level and unstable ratings leave little room to detect change; case difficulty should also be calibrated, and a harder cervical case developed.

### Adherence and Data Completeness

Engagement was substantially higher than in our previous pilot trial, in which only 4 of 23 students allocated to the LLM arm used the model actively [12]. The program in this study was restructured into fewer, thematically related cases with a fixed number of attempts. This suggests that curricular integration, rather than the tool itself, is the main determinant of uptake, consistent with evidence that digital educational tools are underused when they are not embedded in the surrounding curriculum [9,10]. Completion nonetheless differed between cases, with the cervical case completed on both attempts by only 63% of students. Because case order and timing were fixed (lumbar, cervical, thoracic), lower completion may reflect the position of the case in the academic calendar rather than its content. A definitive trial should therefore record completion automatically, with attempt-level time stamps, and should never code a noncompleted attempt as a score of 0, because doing so conflates engagement with performance and, as the sensitivity analysis showed, can inflate validity estimates.

Missing data for secondary measures clustered in the third rotation. Because clinical reasoning competence changes as students progress through successive placements [28], this clustering is not ignorable: it is confounded with the timing of data collection within the academic year. Satisfaction data were missing for all students in the third rotation (overall availability, 56%), which suggests that the questionnaire was not administered in that rotation.

### Acceptability

Mean satisfaction was moderate (6.0 of 10) and lower than the satisfaction reported in our previous pilot trial (median 77 of 100) [12], although the 2 samples and the conditions of administration differ. Satisfaction was not clearly associated with LCJR performance, whereas self-evaluated competence showed a moderate association (ρ=0.49), although with a wide CI. These findings caution against using satisfaction as a proxy for performance. Usability and satisfaction studies of LLM-based simulation often treat positive user experience as a marker of success [29]; our data support reporting acceptability and objectively measured performance as distinct outcomes.

### Implications for a Definitive Trial

On the basis of these findings, we propose the following progression criteria for the next stage, to be specified in advance: at least 80% of students completing all case attempts; at least 90% data completeness for primary and key secondary outcomes in every rotation; LIA case-level internal consistency of at least .70 with no case showing ceiling effects in more than 15% of students; independent, blinded LCJR scoring; and, if LIA scores are to be used as an outcome, agreement between model-generated and human ratings of the same transcripts (eg, ICC ≥.75). Assuming these conditions are met, a 2-arm trial with approximately 39 students per group (78 in total, before allowance for attrition) would provide 80% power to detect a 5-point difference in the LCJR under a conservative variance estimate; adjustment for baseline GPA in an analysis of covariance could reduce this requirement. Because students are taught in groups and rotations, a cluster-randomized or stepped-wedge design across rotations or institutions should be considered to limit contamination between arms.

### Limitations

This study has a single-arm, noncomparative design and cannot speak to the effectiveness of the intervention. The sample of 27 students came from a single institution, and it is the intervention arm of a larger quasi-experimental study, so the 2 reports are not independent. Model-generated and human ratings were obtained in different encounters (virtual vs real patient), so rater effects could not be separated from context effects, and the model was asked to use a 0-4 scale rather than the 1-4 levels of the rubric; agreement between ChatGPT and human raters on the same transcripts was not evaluated, and the reliability of model-generated ratings could be estimated only among completers of each case. The LCJR was developed in nursing, and its validity for physical therapy students has not been established. The GPA-LCJR correlation, the most striking finding in the dataset, is based on 22 of 27 students, and the LCJR was scored by a single rater, the course instructor, so interrater reliability could not be assessed and rater knowledge of students’ academic performance cannot be excluded. Several correlations and paired comparisons were performed without correction for multiplicity. Finally, satisfaction data were available for only about half of the sample, and the LLM used is a general-purpose commercial model whose behavior may change across versions, which limits reproducibility.

### Future Research

Beyond resolving the operational issues identified above, future work should test whether LLM-generated ratings agree with human ratings of the same transcripts and whether their validity improves with explicit behavioral anchors or human scoring, and whether performance in the laboratory predicts clinical judgment over multiple real patient encounters as students accumulate clinical experience. The strong association between academic performance and clinical judgment also raises the question of whether students with lower academic performance benefit differentially from structured LLM-based practice, which would inform whether such tools should be targeted rather than deployed uniformly. Finally, brief interviews or surveys with students and faculty could identify the mechanisms behind case-specific noncompletion and the clustering of missing data in the third rotation.

### Conclusions

A structured LLM-based clinical reasoning laboratory could be delivered within undergraduate physical therapy education with greater engagement than an earlier, less structured approach, but clinical judgment ratings generated by ChatGPT did not reflect those of human assessors with real patients. LLM-generated scores should therefore not be used as outcome or assessment measures until their validity against human raters has been established. Case-specific noncompletion, nonrandom missing data, uneven case difficulty, and the near-perfect association between academic grades and the external criterion are additional, correctable targets before the intervention is evaluated in an adequately powered randomized controlled trial.

## Data Availability

The pseudonymized dataset, data dictionary, and analysis code for this study and its companion quasi-experimental study are available in the Zenodo repository under a CC BY 4.0 license (https://doi.org/10.5281/zenodo.23158824).

## Acknowledgments

We thank all the students who participated in this study. During the preparation of this manuscript, the authors used generative artificial intelligence tools (Claude, Anthropic) for language editing, restructuring of the text, drafting of the statistical analysis code, data cleaning, preparation of figures, and checking of reference formatting, under full human supervision. The authors reviewed and verified all content, including every reference, and take full responsibility for the manuscript. This use is distinct from the use of ChatGPT as part of the educational intervention.

## Authors’ Contributions

ALC: methodology, validation, formal analysis, resources, data curation, writing – original draft, and writing – review & editing. APG: conceptualization, methodology, validation, investigation, resources, writing – original draft, and writing – review & editing. ASV: methodology, validation, investigation, writing – original draft, and writing – review & editing. SDB: methodology, validation, writing – original draft, and writing – review & editing. RFP: conceptualization, methodology, investigation, resources, writing – original draft, writing – review & editing, visualization, supervision, project administration, and funding acquisition.

## Abbreviations

CONSORT: Consolidated Standards of Reporting Trials
GPA: grade point average
ICC: intraclass correlation coefficient
ICF: International Classification of Functioning, Disability and Health
LCJR: Lasater Clinical Judgment Rubric
LIA: Laboratorio de Inteligencia Artificial (artificial intelligence–based clinical reasoning laboratory)
LLM: large language model

## Multimedia Appendix 1

CONSORT checklist for pilot and feasibility trials (adapted for a single-arm design) and CONSORT-EHEALTH (V1.6.1) form.

## Multimedia Appendix 2

Configuration prompt and example interaction for the lumbar virtual patient case.

## Multimedia Appendix 3

Ten-dimension version of the Lasater Clinical Judgment Rubric used by human assessors and by ChatGPT (English translation of the Spanish version).

## Multimedia Appendix 4

Spearman correlation matrix across all instruments.

