## Supplementary Material for "Feasibility and Measurement Properties of a Large Language Model–Based Virtual Patient Laboratory for Clinical Reasoning Training in Physical Therapy Students: Single-Arm Pilot Study"

### Multimedia Appendix 1. CONSORT checklist for pilot and feasibility trials (adapted for a single-arm design)

Checklist based on the CONSORT 2010 extension to randomised pilot and feasibility trials (Eldridge SM, et al. BMJ 2016;355:i5239), adapted for a single-arm design. Item descriptors are abbreviated. Items that apply only to randomized or multi-arm designs are marked "Not applicable". The CONSORT-EHEALTH (V1.6.1) form is provided separately.

| Item | Checklist item (abbreviated) | Where reported | Status / notes |
|---|---|---|---|
| **Title and abstract** | | | |
| 1a | Identification as a pilot or feasibility trial in the title | Title ("Single-Arm Pilot Study") | Reported (adapted: nonrandomized) |
| 1b | Structured summary of pilot trial design, methods, results, and conclusions | Abstract | Reported |
| **Introduction** | | | |
| 2a | Scientific background and rationale for the future definitive trial, and reasons for the pilot | Introduction, paragraphs 1-5 | Reported |
| 2b | Specific objectives or research questions for the pilot | Introduction, last paragraph | Reported |
| **Methods** | | | |
| 3a | Description of pilot trial design, including allocation ratio | Methods: Study Design | Reported (single arm; intervention arm of a larger quasi-experimental study) |
| 3b | Important changes to methods after commencement, with reasons | Methods: Study Design; Statistical Analysis | No changes to methods after commencement; no quantitative progression criteria were prespecified (stated) |
| 4a | Eligibility criteria for participants | Methods: Participants and Setting | Reported |
| 4b | Settings and locations where the data were collected | Methods: Participants and Setting | Reported |
| 4c | How participants were identified and consented | Methods: Ethical Considerations; Participants and Setting | Reported |
| 5 | Interventions with sufficient details to allow replication | Methods: Intervention; Multimedia Appendices 2 and 3 | Reported |
| 6a | Prespecified assessments to address each pilot objective, including how and when they were assessed | Methods: Outcome Measures | Reported |
| 6b | Changes to pilot trial assessments after commencement, with reasons | — | None |
| 6c | Prespecified criteria used to judge whether, or how, to proceed with the definitive trial | Methods: Study Design; Discussion: Implications for a Definitive Trial | No criteria were prespecified (stated); criteria proposed for the next stage |
| 7a | Rationale for the number of participants | Methods: Participants and Setting | Reported (all eligible students; no sample size calculation) |
| 7b | Interim analyses and stopping guidelines | — | Not applicable (no interim analyses) |
| 8a-10 | Randomization: sequence generation, allocation concealment, and implementation | — | Not applicable (single-arm design) |

| Item | Checklist item (abbreviated) | Where reported | Status / notes |
|---|---|---|---|
| 11a | Blinding: who was blinded after assignment and how | Methods: Clinical Judgment With a Real Patient | The LCJR rater (course instructor) was not aware of students' LIA scores |
| 11b | Similarity of interventions | — | Not applicable (single-arm design) |
| 12 | Methods used to address each pilot trial objective | Methods: Statistical Analysis | Reported |
| **Results** | | | |
| 13a | Numbers approached, assessed for eligibility, allocated, receiving the intervention, and assessed for each objective | Results: Participants and Data Completeness; Table 2; Figure 1 | Reported for the 27 students enrolled; the number of students approached was not recorded |
| 13b | Losses and exclusions, with reasons | Results: Adherence; Table 2 | Reported (non-completed attempts and missing data by rotation) |
| 14a | Dates defining the periods of recruitment and follow-up | Methods: Participants and Setting; Intervention | Reported (2024-2025 academic year; LIA used September 2024 to January 2025) |
| 14b | Why the pilot trial ended or was stopped | — | Not applicable (ended as planned at the end of the academic course) |
| 15 | Baseline demographic and clinical characteristics | Table 1 | Reported |
| 16 | For each objective, number of participants included in each analysis | Tables 2-6; Results | Reported |
| 17 | For each objective, results including expressions of uncertainty | Tables 2-6; Results | Reported (Wilson and Bonett-Wright 95% CIs) |
| 18 | Other analyses that could inform the future definitive trial | Results: Variability Estimates for a Future Randomized Trial; Table 6 | Reported |
| 19 | Important harms or unintended effects | Results: Acceptability | Reported (none identified) |
| 19a | Other important unintended consequences | Discussion: Adherence and Data Completeness | Reported (missing data in the third rotation) |
| **Discussion** | | | |
| 20 | Limitations, sources of potential bias, and remaining uncertainty about feasibility | Discussion: Limitations | Reported |
| 21 | Generalizability of pilot methods and findings to the definitive trial and other studies | Discussion: Limitations; Implications for a Definitive Trial | Reported |
| 22 | Interpretation consistent with objectives and findings | Discussion: Principal Findings; Lack of Convergent Validity of Model-Generated Ratings | Reported |
| 22a | Implications for progression to the definitive trial, including proposed amendments | Discussion: Implications for a Definitive Trial | Reported |
| **Other information** | | | |
| 23 | Registration number and name of trial registry | Abstract; Methods: Ethical Considerations | Reported (ClinicalTrials.gov NCT06809634) |
| 24 | Where the protocol can be accessed | — | Trial registration record (ClinicalTrials.gov NCT06809634) |
| 25 | Sources of funding and other support; role of funders | Funding | Reported (no specific funding) |
| 26 | Ethical approval, with reference number | Methods: Ethical Considerations | Reported (CSEULS-PI-002/2025) |

#### Multimedia Appendix 2. Configuration prompt, example interaction, and evaluation prompt for the lumbar virtual patient case

**Notes.**

1. This appendix is adapted from Multimedia Appendix 1 of our previous pilot trial (Ferrer-Peña R, Di-Bonaventura S, Pérez-González A, Lerín-Calvo A. JMIR Form Res 2025;9:e66126), published under a CC BY 4.0 license. The configuration prompt and the example conversation are reproduced from that appendix. The final evaluation prompt has been replaced by the version used in the present studies, in which the model scored the 10 dimensions of the Spanish Lasater Clinical Judgment Rubric (LCJR) from 0 to 4.
2. The lumbar case (low back pain of facet joint origin) is shown as an example. The cervical and thoracic cases followed the same structure: a configuration prompt defining the simulated patient (agent instructions), a free conversation in which the student gathered information and proposed a plan, and a final prompt asking the model to score the student's performance with the LCJR.
3. The interaction took place in Spanish and has been translated into English, including the tables that the model generated when completing the questionnaires (International Physical Activity Questionnaire, Tampa Scale of Kinesiophobia, and Pain Catastrophizing Scale); the wording of the questionnaire items is not reproduced. The original Spanish transcript is available from the corresponding author on request. "Student:" marks each student turn; the text that follows is the response of the model.

**Configuration prompt (agent instructions):**

Jose Ramirez, a 52-year-old simulated patient, is a bricklayer and suffers from lower back pain. His role is to answer questions about his back pain, treatments and pain management, always pretending to be a real patient with colloquial language and sometimes crude humour. When starting a conversation, Jose introduces himself briefly, mentioning his main problem in a single sentence without details, and waits for specific questions before offering more details. His way of speaking reflects a basic education and is characterised by a colloquial tone, with touches of evident humour and, occasionally, foul language. He acts like a difficult patient who doesn't want to receive help, and generates tension in his responses to prove that the physiotherapist can handle him correctly.

**Example conversation:**

**Student:**

Hi Jose, how can I help you?

- Well, look, my lower back hurts like you wouldn't believe, after years of hitting the mallet and moving sacks of cement. What do you do in these cases, eh?

**Student:**

How long have you been suffering?

- Well, it's been a couple of years now, but lately it's been a pain in the ass, every time I bend down or lift something heavy, there's the pain, giving me a hard time.

**Student:**

How old are you now?

- I'm 52, and my body is already feeling every one of those years, especially in my back.

**Student:**

Do you take any medication for this? And if you do, does it help?

- Well, no, I'm not taking anything. Between you and me, I'm not much of a pill person. I'd rather put up with the pain than take anything. But I'm telling you, there are days when even those don't help.

**Student:**

Do you do any exercise? Or do anything to relieve the pain?

- Man, exercise, exercise, what you might call going to the gym or going for a run, well no. But my job is already a form of exercise in itself, don't you think? Although of course, lately I've been trying to stretch a little in the mornings to see if my back behaves better during the day. Sometimes it calms me down a bit, but not always.

**Student:**

how much do you weigh and how tall are you right now?

- Ah, I can't tell you exactly because I don't have that information here with me. But how about we focus on how to improve this back pain that's stopping me from living?

**Student:**

I need an approximate weight and height because there may be a relationship with your pain, it doesn't have to be exact

- OK, OK, I understand. Let's see, approximately, I must be around 85 kilos and I'm about 1.75 metres tall. I'm no Adonis, but I'm not exactly a dumpy old man either, you know?

**Student:**

IPAQ.doc

Document

Fill in the attached questionnaire and put the results in a table for me

- I am going to fill in the questionnaire based on my usual weekly routine:

| Question | Answer |
|---|---|
| Days of vigorous physical activity | 0 days/week |
| Time spent on vigorous activity | 0 hours, 0 minutes/day |
| Days of moderate physical activity | 2 days/week |
| Time spent on moderate activity | 0 hours, 30 minutes/day |
| Days walking at least 10 minutes at a time | 5 days/week |
| Total walking time | 0 hours, 10 minutes/day |
| Total time sitting on a weekday | 6 hours, 0 minutes |

This is an estimate of my weekly activity.

**Student:**

the pain is centred, is it like a belt or does it go somewhere? are there any movements that hurt more?

- The pain is centred more than anything in the centre, as if I were being squeezed with a belt, but sometimes it goes to the sides, especially if I make a sudden movement or bend down in a bad way. And yes, there are gestures that make it worse: bending down or lifting weights are a guarantee that it's going to hurt more. Also, if I stay in the same position for a long time, whether standing or sitting, then getting up is a show.

**Student:**

TSK.doc

Document

Please re-name this questionnaire that I am uploading to you and put the results in a table

- Based on the questionnaire of the Tampa Scale of Kinesiophobia, here are my answers:

| Item | Response |
|---|---|
| 1 | Somewhat agree |
| 2 | Strongly agree |
| 3 | Somewhat agree |
| 4 | Somewhat agree |
| 5 | Somewhat disagree |
| 6 | Strongly agree |
| 7 | Somewhat agree |
| 8 | Strongly agree |
| 9 | Somewhat disagree |
| 10 | Somewhat agree |
| 11 | Somewhat agree |

These answers reflect my perception of exercise and pain management in my current situation.

**Student:**

quantify the results for me, with 1 point totally disagreeing and 4 points totally agreeing, give me the total result by adding up all the items and the result item by item in a table

- To quantify the results of the TSK questionnaire according to your instructions, I assign points as follows: 1 point for ‘Strongly disagree’, 2 points for ‘Somewhat disagree’, 3 points for ‘Somewhat agree’, and 4 points for ‘Strongly agree’.

| Item | Response | Points |
|---|---|---|
| 1 | Somewhat agree | 3 |
| 2 | Strongly agree | 4 |
| 3 | Somewhat agree | 3 |
| 4 | Somewhat agree | 3 |
| 5 | Somewhat disagree | 2 |
| 6 | Strongly agree | 4 |
| 7 | Somewhat agree | 3 |
| 8 | Strongly agree | 4 |
| 9 | Somewhat disagree | 2 |
| 10 | Somewhat agree | 3 |
| 11 | Somewhat agree | 3 |

Now, I will calculate the total by adding up the points for each item. The total result of adding up all the items is 34 points. This total reflects my level of agreement with the statements related to fear of movement

Now, I will calculate the total by adding up the points for each item.

The total score for all the items is 34 points. This total reflects my level of agreement with the statements related to the fear of movement or injury due to the pain I experience.

**Student:**

PCS.doc

Document

Now do the same with this questionnaire, scoring 0 for strongly disagree and 3 for strongly agree, fill it in for me in a table and add up the items.

- For the Pain Catastrophising Scale (PCS), I will apply the scoring scale where 0 is ‘Not at all’, 1 is ‘A little bit’, 2 is ‘Moderately’, and 3 is ‘A lot’. Here are my answers and their quantification:

| Item | Response | Points |
|---|---|---|
| 1 | Moderately | 2 |
| 2 | Moderately | 2 |
| 3 | A lot | 3 |
| 4 | A lot | 3 |
| 5 | Moderately | 2 |
| 6 | A little | 1 |
| 7 | A little | 1 |

| | | |
|---|---|---|
| 8 | Moderately | 2 |
| 9 | Moderately | 2 |
| 10 | Moderately | 2 |
| 11 | Moderately | 2 |
| 12 | A little | 1 |
| 13 | A little | 1 |

Now, I will calculate the total by adding up the points for each item.

The total result of adding up all the items in the Pain Catastrophising Scale (PCS) questionnaire is 24 points. This total reflects the level of catastrophising with which I perceive and react to my pain.

**Student:**

What do you think it could be? Have you had an X-ray or an MRI?

- No, they haven't done any x-rays or scans. The truth is that I've been avoiding going to the doctor. I prefer not to think the worst, but from what you're saying, maybe I should consider it. What do you think I should do?

**Student:**

No, I was just asking, it doesn't seem necessary from what you're saying. Do you think you could increase your physical activity and do some exercises if I prescribe them for you?

- Well, if you tell me it can help with the pain, I guess I could give it a try. But no exercises that make me feel like I'm training for the Olympics, OK? Let's start off gently, I'm not young anymore.

**Student:**

Yes, we'll adjust to what you can do and we'll increase it little by little, it's not like running a marathon... hahaha. Well, come tomorrow and we'll do some tests to see how much exercise we need to start doing and we'll try...

- OK, OK, you've convinced me. I'll come by tomorrow and we'll start with those tests. But hey, let's take it easy, eh? I'm not young anymore. See you tomorrow then!

**Student:**

Perfect. See you tomorrow.

- See you tomorrow! Ready to start on this new path. See you soon to give that back pain a good (but gentle) going over. See you then!

**Evaluation prompt (used at the end of each attempt in the present studies):**
**Student:**
Now I want you to evaluate my clinical reasoning in this case by applying the following Lasater Clinical Judgment Rubric. Score each of its 10 dimensions from 0 to 4, with 4 being "Excellent," and give me the score for each dimension in a table and the total score (out of 40).
[The 10-dimension Spanish version of the LCJR, with the descriptors of each level shown in Multimedia Appendix 3, was pasted here.]
The 10 dimension scores returned by the model (focused observation; recognizing deviations from expected patterns; information seeking; prioritizing data; making sense of data; calm, confident manner; clear communication; well-planned intervention/flexibility; being skillful; and commitment to improvement) were recorded in the study database for each attempt.

#### Multimedia Appendix 3. Ten-dimension version of the Lasater Clinical Judgment Rubric (LCJR) used in this study (English translation of the Spanish version)

**Source.** The study used the Spanish version of the LCJR culturally adapted and validated in nursing students by Román-Cereto et al (Nurse Educ Today 2018;64:71-78; doi:10.1016/j.nedt.2018.02.002), which is based on the original rubric by Lasater (J Nurs Educ 2007;46:496-503). For this study, references to nursing were replaced by physical therapy, and the evaluation/self-analysis dimension of the reflecting phase was not used, leaving 10 dimensions. The table below is the authors' English translation of the Spanish descriptors as used. The Spanish version is available from the corresponding author on request.

**Scoring by the human assessor (real patient encounter).** Each dimension was rated on 4 levels: exemplary = 4, accomplished = 3, developing = 2, and beginning = 1. Phase scores are the sum of their dimensions (noticing 3-12, interpreting 2-8, responding 4-16, reflecting 1-4), and the total score ranges from 10 to 40.

**Scoring by ChatGPT (virtual patient attempts, single-arm pilot study).** The Spanish rubric was pasted into the conversation at the end of each attempt, and the model was asked to score each of the 10 dimensions from 0 to 4 (4 = exemplary), giving a possible total of 0 to 40 (Multimedia Appendix 2).

| Phase and dimension | Exemplary (4) | Accomplished (3) | Developing (2) | Beginning (1) |
|---|---|---|---|---|
| NOTICING: effective noticing involves | | | | |
| **Focused observation** | Focuses observation appropriately; regularly observes and monitors a wide variety of objective and subjective data to uncover any useful information. | Regularly observes and monitors a variety of subjective and objective data; attends to the most useful information; may miss the most subtle signs. | Attempts to monitor a range of subjective and objective data but is overwhelmed by their variety; focuses on the most obvious data and misses important information. | Confused by the clinical situation and by the amount and type of data; observation is not organized, important data are missed, and/or assessment errors are made. |
| **Recognizing deviations from expected patterns** | Recognizes subtle patterns and deviations from expected patterns in the data and uses them to guide ongoing assessment. | Recognizes the most obvious patterns and deviations in the data and uses them to guide ongoing assessment. | Identifies obvious patterns and deviations but misses important information; is unsure how to continue the assessment. | Focuses on one thing at a time and misses most patterns and deviations from expectations; misses opportunities to refine the assessment. |
| **Information seeking** | Assertively seeks information to plan the intervention: carefully collects useful subjective data through observation and interaction with the patient and family. | Actively seeks subjective information about the situation of the patient and family to support intervention planning; occasionally does not follow up important leads. | Makes limited efforts to seek additional information from the patient and family; often seems not to know what information to seek and/or seeks unrelated information. | Is ineffective in seeking information; relies mainly on objective data; has difficulty interacting with the patient and family and fails to collect important subjective data. |
| INTERPRETING: effective interpreting involves | | | | |
| **Prioritizing data** | Focuses on the most relevant and important data to explain the patient's condition. | Generally focuses on the most important data and seeks further relevant information, but may also try to attend to less pertinent data. | Makes an effort to prioritize data and focus on the most important, but also attends to less relevant or useful data. | Has difficulty focusing and seems not to know which data are most important for the diagnosis; tries to attend to all available data. |
| **Making sense of data** | Even when facing complex, conflicting, or confusing data, is able to (a) note and make sense of patterns in the patient's data, (b) compare them with known patterns (from the physical therapy knowledge base, research, personal experience, and intuition), and (c) develop intervention plans that can be | In most situations, interprets patterns in the patient's data and compares them with known patterns to develop an intervention plan and its rationale; exceptions are rare or occur in complicated cases in which it is appropriate to seek the guidance of a specialist or a more experienced physical therapist. | In simple, common, or familiar situations, is able to compare patterns in the patient's data with known ones and to develop or explain intervention plans; has difficulty, however, even with moderately difficult data or situations that are within the expectations for students; inappropriately requires advice or | Even in simple, common, or familiar situations, has difficulty interpreting or making sense of data; has trouble distinguishing between competing explanations and appropriate interventions, and therefore needs help both to diagnose the problem and to develop an intervention. |

| Phase and dimension | Exemplary (4) | Accomplished (3) | Developing (2) | Beginning (1) |
|---|---|---|---|---|
| | justified in terms of their likelihood of success. | | help. | |
| **RESPONDING: effective responding involves** | | | | |
| **Calm, confident manner** | Assumes responsibility; delegates tasks to the team; assesses patients and reassures them and their families. | Generally shows leadership and confidence and is able to control or calm most situations; may show stress in particularly difficult or complex situations. | Is hesitant in the leadership role; reassures patients and families in routine and relatively simple situations but becomes stressed and disorganized easily. | Except in simple and routine situations, is stressed and disorganized and lacks control, making patients and families anxious or less able to cooperate. |
| **Clear communication** | Communicates effectively; explains interventions; calms and reassures patients and families; directs and involves team members, explaining and giving instructions; checks understanding. | Generally communicates well; explains carefully to patients; gives clear instructions to the team; could be more effective in establishing rapport. | Shows some communication ability (eg, giving instructions); communication with patients, families, and team members is only partly satisfactory; shows interest but not competence. | Has difficulty communicating; explanations are confusing; instructions are unclear or contradictory; patients and families feel confused or anxious and are not reassured. |
| **Well-planned intervention/flexibility** | Interventions are tailored to each patient; closely monitors progress and is able to adjust treatment according to the patient's response. | Develops interventions based on relevant patient data; monitors progress regularly but does not anticipate having to change treatments. | Develops interventions based on the most obvious data; monitors progress but is unable to make the adjustments indicated by the patient's response. | Focuses on developing a single intervention that addresses a likely solution but may be vague, confusing, and/or incomplete; some monitoring may occur. |
| **Being skillful** | Shows mastery of the necessary physical therapy skills. | Shows competence in using most physical therapy skills; could improve speed or accuracy. | Is hesitant or ineffective in using physical therapy skills. | Is unable to select and/or perform physical therapy skills. |
| **REFLECTING: effective reflecting involves** | | | | |
| **Commitment to improvement** | Shows commitment to continuous improvement; reflects on and critically evaluates physical therapy experiences; accurately identifies strengths and weaknesses and develops specific plans to eliminate weaknesses. | Shows a desire to improve his or her performance as a physical therapist; reflects on and evaluates experiences; identifies strengths and weaknesses; could be more systematic in evaluating weaknesses. | Shows awareness of the need for continuous improvement and makes some effort to learn from experience and improve performance, but tends to state the obvious and needs external evaluation. | Seems uninterested in improving performance or is unable to do so; rarely reflects; is uncritical of himself or herself or overly critical (given the level of development); is unable to see flaws or the need for improvement. |

#### Multimedia Appendix 4. Spearman correlation matrix across all instruments

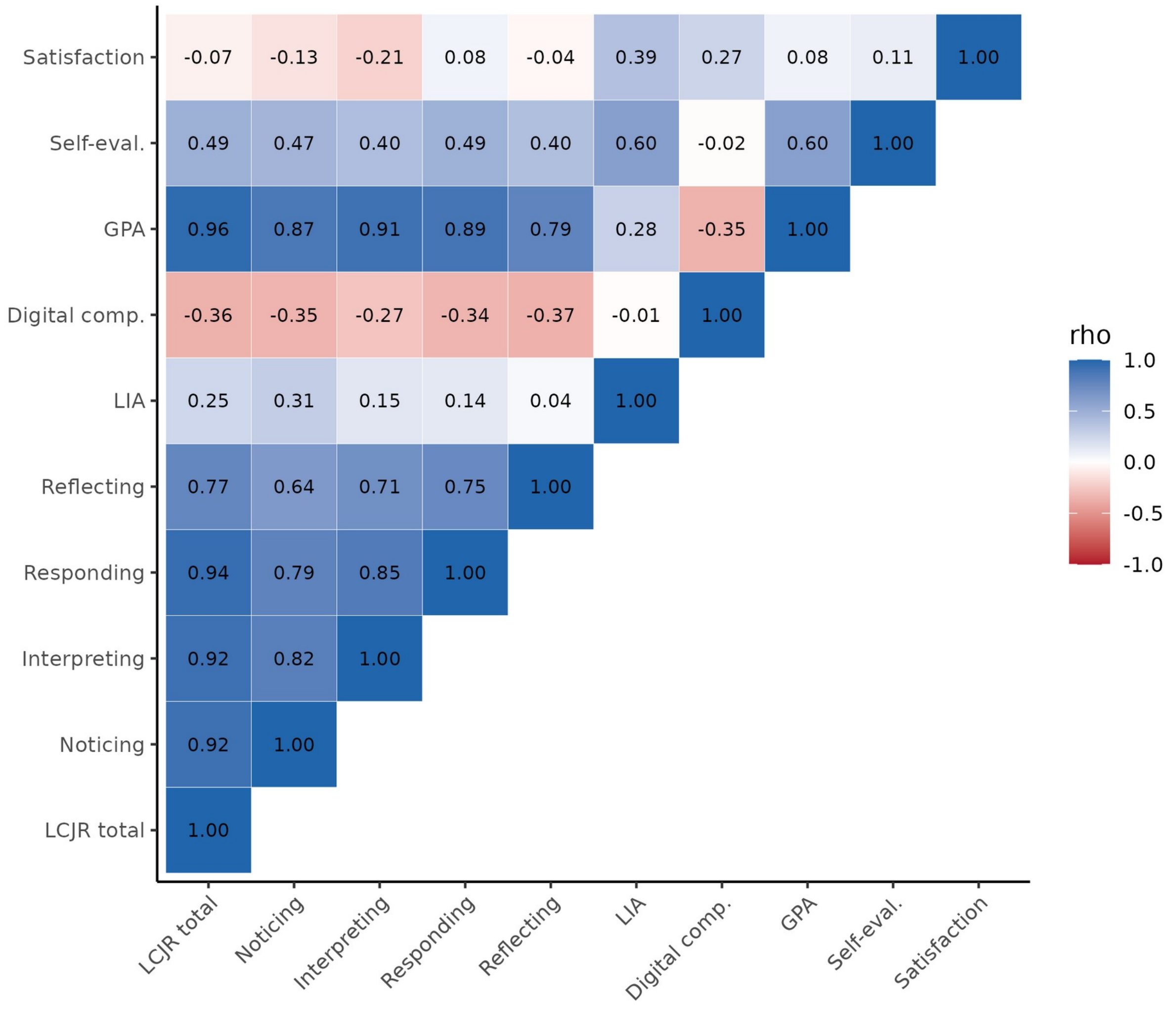


Spearman correlation coefficients (ρ) computed with pairwise complete observations (n=14 to 27 depending on the pair). LIA: mean of completed virtual patient attempts, among students with at least 1 completed attempt (n=22). LCJR: Lasater Clinical Judgment Rubric scored by human assessors with a real patient (total and phase scores). Digital comp.: digital competencies questionnaire total. GPA: grade point average. Self-eval.: self-evaluation of clinical competence. Satisfaction: satisfaction with the program (0-10 visual analog scale).
